# Design and implementation of a global site assessment survey among HIV clinics participating in the International epidemiology Databases to Evaluate AIDS (IeDEA) research consortium

**DOI:** 10.1101/2022.04.25.22274292

**Authors:** Ellen Brazier, Fernanda Maruri, C. William Wester, Beverly Musick, Aimee Freeman, Angela Parcesepe, Stefanie Hossmann, Benedikt Christ, April Kimmel, John Humphrey, Esther Freeman, Leslie A. Enane, Kathryn E. Lancaster, Marie Ballif, Jonathan E. Golub, Denis Nash, Stephany N. Duda, International epidemiology Databases to Evaluate AIDS (IeDEA) consortium

## Abstract

**Introduction:** Timely descriptions of HIV service characteristics and their evolution over time across diverse settings are important for monitoring the scale-up of evidence-based program strategies, understanding the implementation landscape, and examining service delivery factors that influence HIV care outcomes.

**Methods:** The International epidemiology Databases to Evaluate AIDS (IeDEA) consortium undertakes periodic cross-sectional surveys on service availability and care at participating HIV treatment sites to characterize trends and inform the scientific agenda for HIV care and implementation science communities. IeDEA’s 2020 general site assessment survey was developed through a consultative, 18-month process that engaged diverse researchers in identifying content from previous surveys that should be retained for longitudinal analyses and in developing expanded and new content to address gaps in the literature. An iterative review process was undertaken to standardize the format of new survey questions and align them with best practices in survey design and measurement and lessons learned through prior IeDEA site assessment surveys.

**Results:** The survey questionnaire developed through this process included eight content domains covered in prior surveys (patient population, staffing and community linkages, HIV testing and diagnosis, new patient care, treatment monitoring and retention, routine HIV care and screening, pharmacy, record-keeping and patient tracing), along with expanded content related to antiretroviral therapy (differentiated service delivery and roll-out of dolutegravir-based regimens); mental health and substance use disorders; care for pregnant/postpartum women and HIV-exposed infants; tuberculosis preventive therapy; and pediatric/adolescent tuberculosis care; and new content related to Kaposi’s sarcoma diagnostics, the impact of COVID-19 on service delivery, and structural barriers to HIV care. The survey was distributed to 238 HIV treatment sites in late 2020, with a 95% response rate.

**Conclusion:** IeDEA’s approach for site survey development approach has broad relevance for HIV research networks and other priority health conditions.

## I. Introduction

HIV care and treatment programs around the world continue to evolve and expand. However, the changing characteristics of HIV services are not well described across countries and global regions. Timely descriptions of the evolution of and variations in the characteristics of HIV services over time are important for documenting the comprehensiveness of care provided and progress in the scale-up of services and evidence-based service delivery strategies, as well as advancing the global HIV community’s implementation science agenda [1]. Such data can enhance understanding of the implementation landscape for HIV care across the globe, while being essential for efforts to identify service delivery factors that are associated with care outcomes of interest among those newly enrolling in HIV care and those engaged in long-term HIV care.

The International epidemiology Database to Evaluate AIDS (IeDEA) consortium (http://www.iedea.org) is a global research consortium of HIV care and treatment sites in seven geographic regions: the Asia-Pacific, the Caribbean, Central, and South America (CCASAnet), North America (NA-ACCORD), and sub-Saharan Africa: Central Africa, East Africa, Southern Africa, and West Africa. Established in 2006, the IeDEA research consortium currently comprises close to 400 HIV care and treatment sites in 44 countries that contribute longitudinal data for more than 2 million patients ever enrolled in HIV care—data that are used in epidemiology research on HIV care and treatment outcomes.

IeDEA undertakes periodic surveys among sites that are active in the consortium in order to document HIV service availability across sites and over time—information that is helpful for understanding patient-level data and examining how patient outcomes are associated with health facility and service delivery characteristics. We herein describe the process and methodology used in developing IeDEA’s 2020 site assessment survey and fielding it across treatment sites within the research consortium.

## II. Materials and methods

### Study aim and objectives

Every three to five years, IeDEA surveys the majority of HIV care and treatment clinics that are active in the consortium to collect information on service provision and practices at participating sites. Designed as a cross-sectional facility-level survey of HIV treatment sites, the objectives of IeDEA’s 2020 consortium-wide survey were to: (1) systematically document facility, program, and service characteristics of IeDEA sites, including selected clinic and service delivery attributes that change over time; (2) collect data on specialized topics related to HIV care that constitute important gaps in the scientific literature; (3) compile preliminary data for future research studies; and (4) collect data to inform the development and implementation of future in-depth topic-specific site assessment surveys, as well as intervention studies.

### Survey development process

The content of IeDEA’s 2020 site assessment survey was developed through a consultative 18-month process (Fig 1) that built on lessons learned through prior IeDEA-wide surveys [2-4], while harnessing the expertise of researchers across IeDEA. As a first step in this process, members of IeDEA’s technical working groups (TWGs) [5] were invited to review the content of the previous general site assessment survey, conducted in 2017, and make recommendations for content that should be retained to facilitate longitudinal analyses of service delivery and practices across IeDEA sites. IeDEA’s TWGs were also asked to identify questions that could be retired because they were no longer relevant or because of concerns with the quality of data generated during the previous survey. In addition, data from the 2017 survey were analyzed to identify response options that should be collapsed or combined to avoid counts too small for meaningful analysis. Through this review process, core domains of the survey questionnaire were defined.

**Fig 1.**
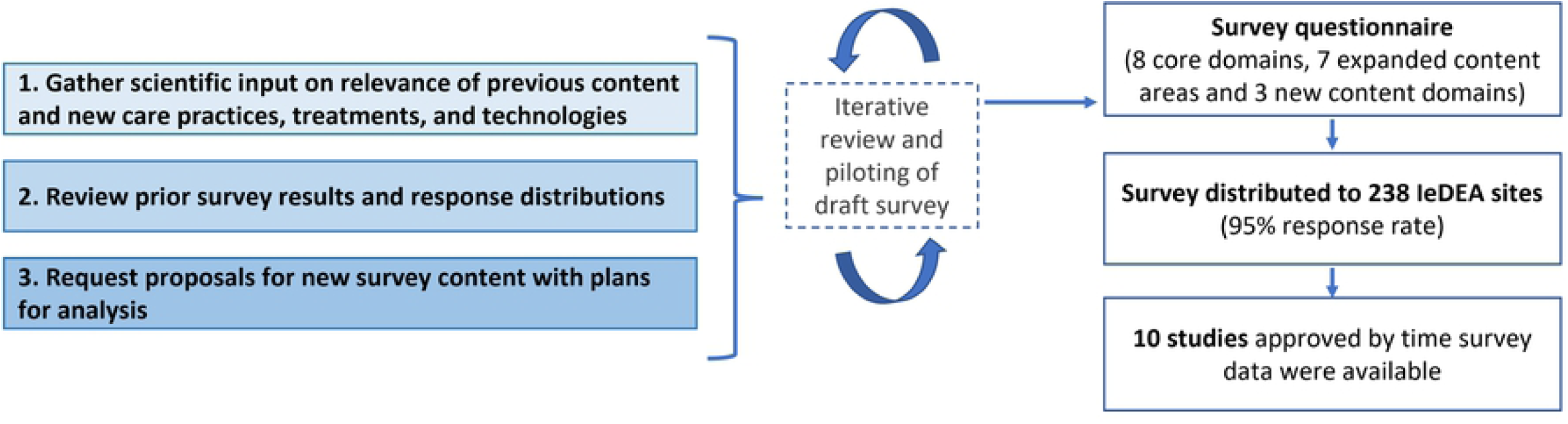
Survey development process (2019-2021)

In addition to reviewing historical content, a complementary proposal-driven process was undertaken for the development of expanded and new survey content. Each of IeDEA’s TWGs were invited to prepare brief research proposals to support the addition of new survey content, articulating focused objectives and proposing specific survey questions to address key gaps in the literature. Investigators proposing new survey content were requested to specify plans for using the data (e.g., as the basis for an analysis that would result in a manuscript and/or as pilot data to inform a more comprehensive research study). An iterative review process was undertaken to standardize the format of new survey content, ensuring that questions conformed with best practices in site-level survey design and measurement developed through prior IeDEA site assessment surveys and used uniform response options, branching logic and skip patterns to help minimize respondent burden. Through this process, expanded content was developed for several domains covered by prior surveys, and several new survey domains were proposed.

While it was originally intended that the survey would explore the status of service delivery at the time of survey implementation, calendar year 2019 was selected as the primary reference period for most survey questions because of disruptions related to the SARS-CoV-2 pandemic in 2020. This pre-COVID-19 reference period was also selected because several of the associated research proposals planned to link site survey data with patient-level data for the pre-pandemic period.

After compiling the survey content as a draft paper questionnaire, the questionnaire was programmed in REDCap (Research Electronic Data Capture)—a web-based software platform for secure data capture in research studies. REDCap provides a user-friendly interface for data capture; audit capabilities for tracking data manipulation; automated export routines for common statistical packages (e.g., R, SAS, Stata, SPSS); and procedures for data integration with external sources [6, 7].

Housed in a local data center hosted at Vanderbilt University Medical Center (VUMC), the REDCap database for the 2020 survey was extensively tested and refined through an iterative review process to ensure that the online survey was streamlined for data entry and easy to navigate. Once the online data capture instruments were completed in mid-2020, the survey was piloted in English at eight sites across four IeDEA regions. The pilot served to identify questions and wording that were not consistently understood, as well as various modifications to improve the survey instructions, response options, skip patterns, and technical content of the questionnaire.

After addressing issues emerging through the pilot, the survey content and REDCap database were finalized in English, and the questionnaire was then translated into French for use in Francophone settings. The translation was completed by a professional translator with specialized expertise in health care, pharmaceuticals, and HIV. The draft translation was then reviewed by IeDEA colleagues in the West Africa region to ensure that it accurately reflected the idiomatic and technical terms predominant in the region. The final text in the French questionnaire was used in a second deployment of the REDCap database for IeDEA-participating sites in Francophone countries.

## III. Results

### Survey content

The survey developed through IeDEA’s consultative 18-month process covered eight core domains included in prior surveys, including patient populations served, clinic staffing, community linkages, HIV testing and diagnostic capacity, care for newly-enrolling patients, antiretroviral therapy (ART) adherence and retention strategies, routine care and screening for HIV patients, pharmacy services, and record-keeping (S1). In addition, it included expanded content in seven thematic areas (differentiated ART service delivery; roll-out of dolutegravir-based regimens; screening and management of mental health and substance use disorders; TB preventive therapy; pediatric and adolescent TB care; and pregnancy and postpartum care for women and HIV exposed infants) and three new content domains (structural barriers to HIV-related care, Kaposi’s sarcoma diagnostics, and the impact of COVID-19 on routine HIV care and treatment).

### Sampling procedures

All sites that were actively contributing longitudinal patient-level data to IeDEA in 2020 were eligible for inclusion in the survey. Sites that were exiting IeDEA in 2020 (n=3) were ineligible for inclusion, along with sites that were part of an interval cohort (n=14) (i.e., sites where patient data reflect systematic assessments at pre-specified and regular intervals, rather routine patient care extracted from medical records). In the Southern Africa region of IeDEA, where more than 200 small clinics contribute data as part of large programmatic cohorts, surveying all clinics was determined to be unfeasible. Accordingly, a systematic, hybrid sampling strategy was used in Southern Africa to select 23 sites from large programmatic cohorts. To facilitate longitudinal analyses of HIV care and service attributes across time, purposive sampling was used to select five sites that had completed prior IeDEA-wide surveys. In addition, to ensure the representativeness of sites selected from large programmatic cohorts, random sampling, stratified by urban vs. rural location in accordance with the distribution of clinics within the cohort, was used to select an additional 18 sites for inclusion in the survey. In addition to these 23 purposively and randomly selected sites, nine sites that are not part of a large programmatic cohort were eligible for inclusion.

The final sample eligible for the survey comprised 238 sites (Fig 2), including all active sites in the Asia-Pacific region (n=52), CCASAnet (n=9), Central Africa (n=21), East Africa (n=74), West Africa (n=14), along with all sites in NA-ACCORD that are not part of an interval cohort (n=36), and 32 sites from the Southern Africa region (15% of all active sites in the region).

**Fig 2:**
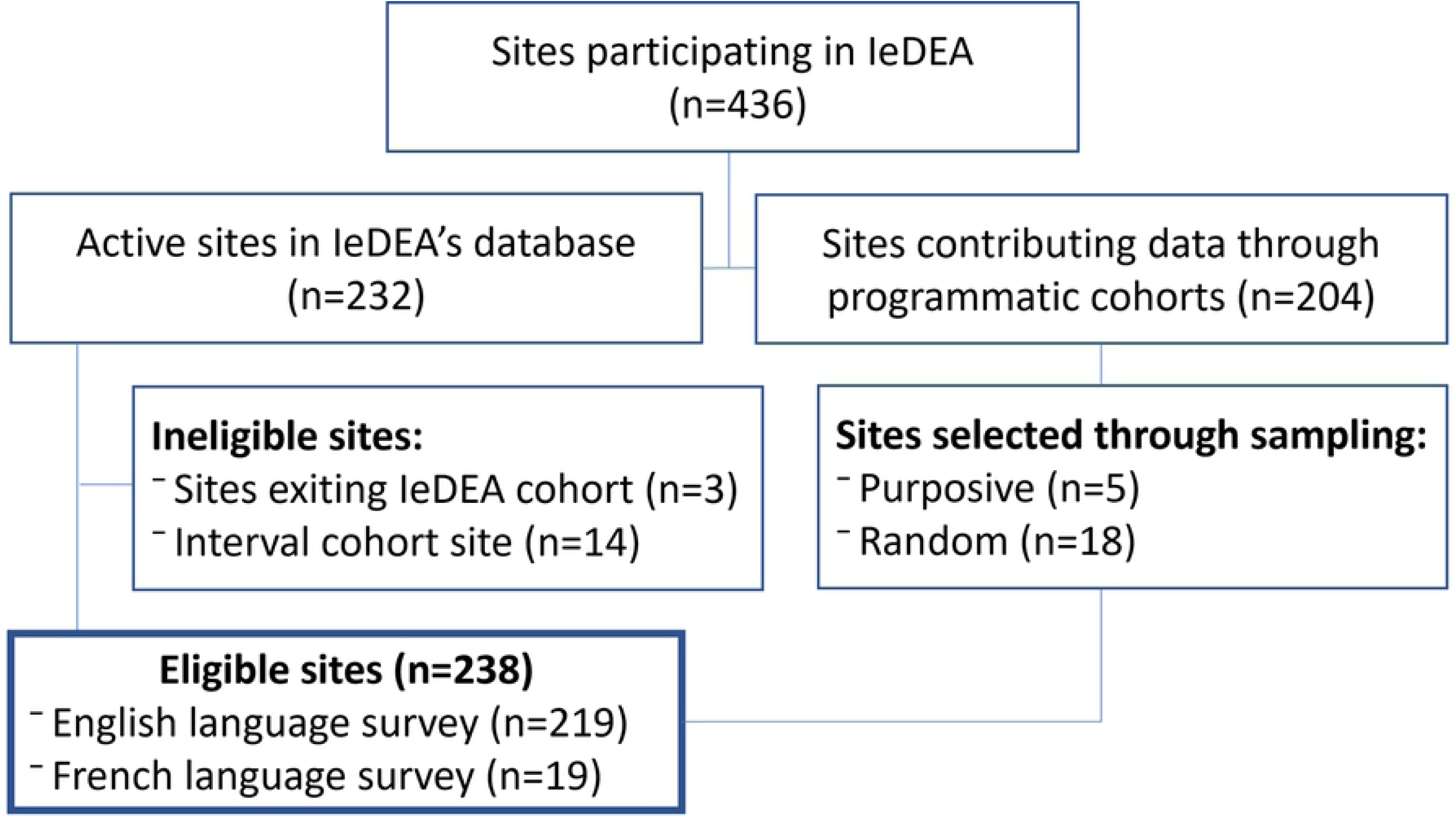
Site inclusion flowchart.

### Ethical review

The conduct of the 2020 IeDEA-wide site assessment survey was reviewed and designated a non-human subjects operational/quality improvement project by the VUMC Institutional Review Board (#200013).

### Survey implementation

Targeted at clinic staff with in-depth knowledge of the care and services provided to adult and pediatric HIV patients, the survey was self-administered in either English or French, using paper forms and online REDCap electronic versions of the questionnaire, depending on country context. Regional data managers affiliated with IeDEA distributed the survey instructions, a PDF of the questionnaire, and unique site-specific survey links to all eligible sites within their region, with the English-language version of the survey launched on September 11, 2020, and the French-language version launched one month later, on October 16, 2020. Sites that had participated in the pilot were asked only to respond to new and revised questions. Throughout the data collection period, data quality checks were performed centrally by regional data managers to identify incomplete surveys and surveys with inconsistent data so that in-country coordinators could follow-up with specific sites, as needed, with reminders and queries to maximize the survey response rate and resolve data quality issues. Surveys completed on paper were entered into the REDCap electronic database by local or regional data managers.

When the survey was closed on March 1, 2021, responses had been received from 227 sites across IeDEA’s seven regions (Fig 3)—an overall response rate of 95% (11 sites did not begin the survey), with 98% of submitted surveys having complete responses (Table 1). Most survey respondents (88%) identified themselves as clinical staff or managers (e.g., clinical officer in charge, clinic manager, or other clinical officer), with 9% of surveys completed by site-level data managers or affiliated research staff and 4% not specified. Almost half (48%) of the surveys were completed by more than one staff (range: 2 to 18), meaning that the survey responses reflected the perspectives of multiple staff and providers involved in HIV-related care.

**Table 1.** 2020 IeDEA-wide site assessment survey response and completion rates.

| Region | Eligible sites<br>n | Survey Response<br>n (%) | Survey Completion<br>n (%) |
| --- | --- | --- | --- |
| Asia Pacific | 52 | 51 (98%) | 51 (100%) |
| CCASAnet | 9 | 9 (100%) | 8 (89%) |
| Central Africa* | 21 | 21 (100%) | 21 (100%) |
| East Africa | 74 | 74 (100%) | 74 (100%) |
| NA-ACCORD | 36 | 30 (83%) | 29 (97%) |
| Southern Africa | 32 | 28 (88%) | 26 (93%) |
| West Africa† | 14 | 14 (100%) | 14 (100%) |
| **Total** | **238** | **227 (95%)** | **223 (98%)** |

\* English version survey was completed by 15 sites, French version survey was completed by 6 sites
† English version survey was completed by one site, French version survey was completed by 13 sites

**Fig 3:**
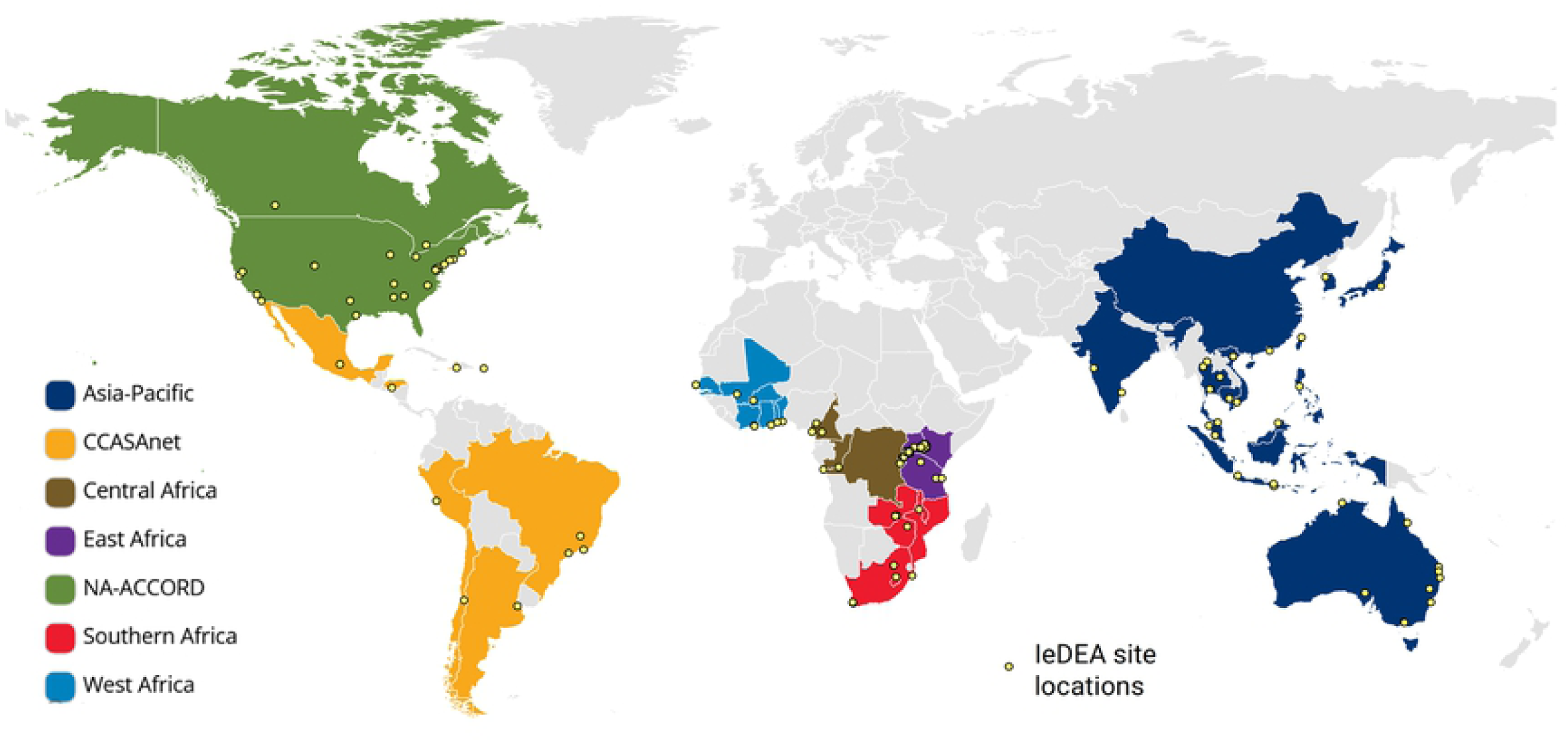
IeDEA regions and locations of responding sites.

### Data management

After the survey closed, additional data quality checks were performed to review data completeness and consistency, and site-level respondents were queried, as needed, to resolve data quality issues. Following the completion of data quality checks in April 2021, data from the English and French databases were merged, and REDCap’s automated export tools were used to extract the data for use in analyses. All survey information, including survey instructions, English and French questionnaires, linked research proposals, and data exports were stored in <u>R</u>esearch <u>O</u>rganization <u>C</u>ollaboration & <u>K</u>nowledge <u>E</u>xchange <u>T</u>oolkit (ROCKET) [8], a HIPAA-compliant web-based tool housed in VUMC’s StarBRITE research portal [9] for dissemination to IeDEA investigators. By June 2021, data had been released for all analyses planned through the survey development process, along with an additional longitudinal analysis (Table 2).

**Table 2.**
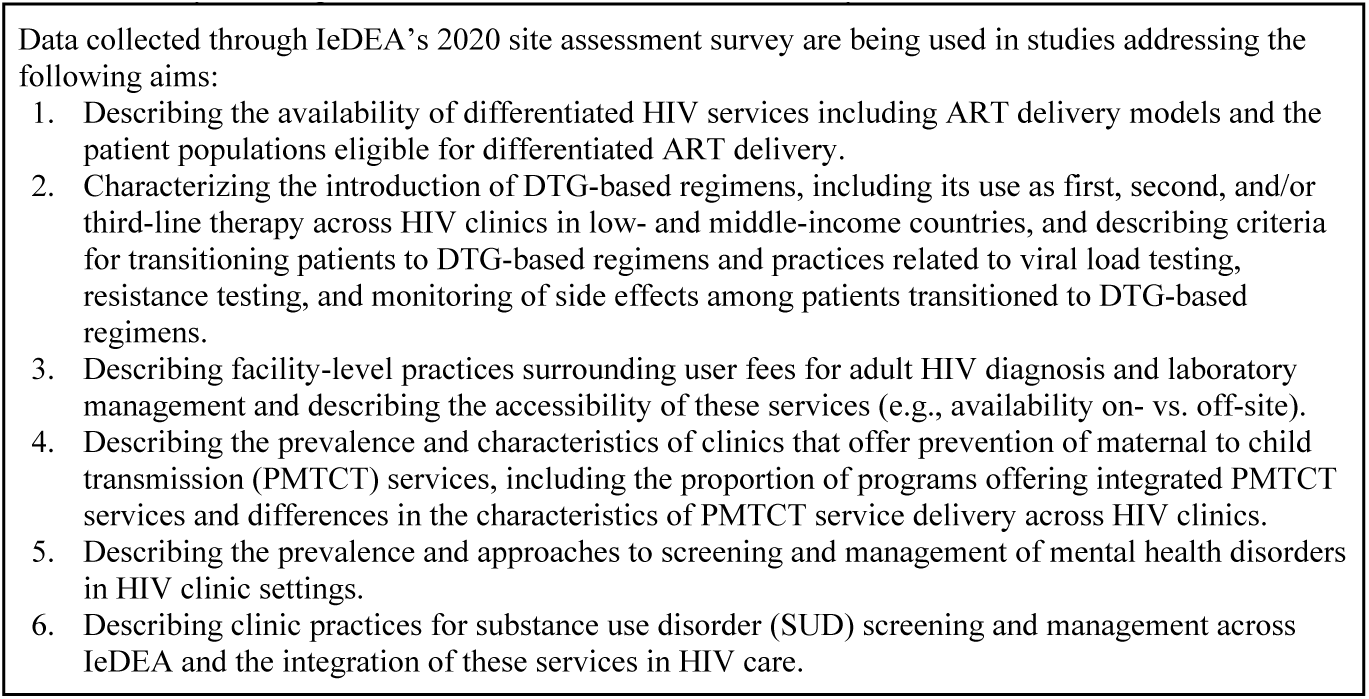
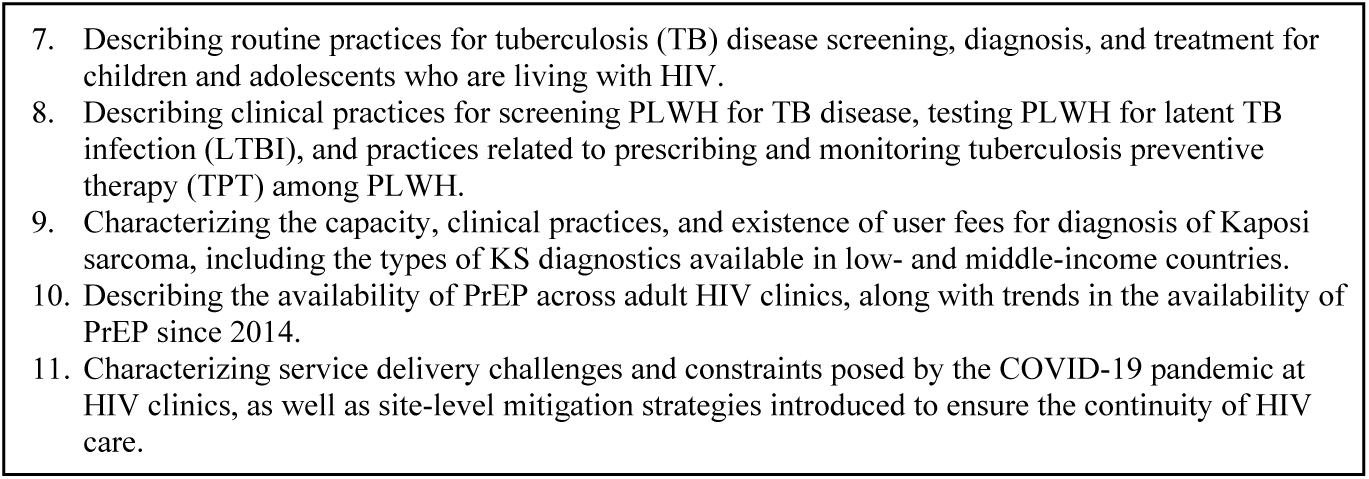
Analyses using the IeDEA 2020 site assessment survey data.

## III. Discussion

IeDEA’s consortium-wide site assessment surveys provide an opportunity to document the evolution of HIV care and treatment for adults and children among a globally diverse cohort of clinics at all levels of the health system, in urban and rural areas and in low-, middle-, and high-income settings. IeDEA’s established relationships with participating sites allow us to collect timely data on the adoption and implementation of new guidance and best practices among real-world service delivery programs in order to track progress and identify implementation gaps. The ability to link this programmatic information with data for patients enrolled in HIV care at participating sites, further allows for investigation of associations between program and policy implementation and patient outcomes of interest.

Important strengths of IeDEA’s 2020 site assessment survey development process include data-driven and proposal-driven approach, which engaged diverse methodological and scientific expertise within the consortium. By starting with an analysis of prior IeDEA-wide survey data, we were able to identify questions and response options that have not provided reliable or useful information previously. In addition, ensuring that new survey content was developed by investigators involved in IeDEA’s TWGs and supported by detailed, aim-driven proposals with specific plans for using the data helped ensure that time and effort expended on data collection led to analyses and manuscripts that address salient and timely questions for the global HIV response. With work in progress for 11 papers addressing gaps in the literature, IeDEA’s 2020 site assessment survey promises to generate more scientific productivity than IeDEA’s prior consortium-wide general site assessment surveys [2-4] and specialized surveys focused on specific patient populations (e.g., pediatrics, pregnant and postpartum women) [10] and areas of service delivery (e.g., tuberculosis, cancer, mental health disorders, etc.) [11-17].

Several limitations of IeDEA’s site assessment survey implementation process are worth noting. First, these surveys rely on voluntary self-report by survey respondents. While the presence or absence of specific services or care components can be captured rather reliably with minimal misclassification, it is also likely that knowledge about HIV care and service delivery varies across respondent types and across sites. Moreover, social desirability bias may result in some respondents reporting practices outlined in relevant norms and guidelines, as opposed to their real world, day-to-day practices. Recall bias is another potential concern, particularly as the survey primarily focused on practices prior to the COVID-19 pandemic and because 2020 was a year with extraordinary challenges for health service delivery around the globe, with many health care facilities introducing changes in practice to mitigate risks associated with the pandemic. It is therefore possible that recall of services and practices prior to the pandemic were inaccurate or incomplete, potentially resulting in either over- or under-reporting of services. While such biases cannot be completely avoided in voluntary self-administered surveys, our high response and survey completion rates, as well as the fact that almost half of the sites involved multiple staff in completing the survey may partially mitigate the impact of these biases.

In addition, IeDEA sites may not be representative of HIV service delivery within some countries and regions, particularly in contexts where IeDEA sites were initially identified because they served a sizable patient population and/or offered advanced levels of care. This limitation notwithstanding, as IeDEA’s 2020 site assessment survey included a large number of sites and settings that have participated in previous IeDEA-wide surveys, it is expected that temporal trends in HIV service delivery among IeDEA sites may be broadly reflective of prevailing trends across other HIV sites that are not part of the consortium [18].

## IV. Conclusion

IeDEA’s consortium-wide site assessments position it to address timely knowledge gaps related to HIV care and treatment service delivery, as well as to substantially contribute to the field of HIV implementation science globally. The collaborative approach used in developing IeDEA’s 2020 global site assessment survey has broad applicability for other large HIV service provider networks and programs, along with research and service provision networks addressing other health conditions, such as tuberculosis and other non-communicable conditions.

## Data Availability

All relevant data are within the manuscript and its Supporting Information files.

## Acknowledgements

The authors would like to thank site investigators, clinicians and data managers, as well as regional data centers who contribute to IeDEA and members of the IeDEA Site Assessment Working Group. A complete list of acknowledgments can be found in the Supporting Information.

